# Excess mortality for care home residents during the first 23 weeks of the COVID-19 pandemic in England: a national cohort study

**DOI:** 10.1101/2020.11.11.20229815

**Authors:** Marcello Morciano, Jonathan Stokes, Evangelos Kontopantelis, Ian Hall, Alex J Turner

**Author notes:** Corresponding Author and Guarantor. The Corresponding Author has the right to grant on behalf of all authors. The guarantor affirms that the manuscript is an honest, accurate, and transparent account of the study being reported; that no important aspects of the study have been omitted.

## Abstract

**Background:** To estimate excess mortality for care home residents during the COVID-19 pandemic in England, exploring associations with care home characteristics.

**Methods:** Daily number of deaths in all residential and nursing homes in England notified to the Care Quality Commission (CQC) from 1^st^ January 2017 to 7^th^ August 2020. Care home level data linked with CQC care home register to identify homes characteristics: client type (over 65s/children and adults), ownership status (for-profit/not-for-profit; branded/independent), and size (small/medium/large).

Excess deaths computed as the difference between observed and predicted deaths using local authority fixed-effect Poisson regressions on pre-pandemic data. Fixed-effect logistic regressions were used to model odds of experiencing COVID-19 suspected/confirmed deaths.

**Findings:** Up to 7^th^ August 2020 there were 29,542 (95%CI: 25,176 to 33,908) excess deaths in all care homes. Excess deaths represented 6.5% (95%CI: 5.5% to 7.4%) of all care home beds, higher in nursing (8.4%) than residential (4.6%) homes. 64.7% (95%CI: 56.4% to 76.0%) of the excess deaths were confirmed/suspected COVID-19. Almost all excess deaths were recorded in the quarter (27.4%) of homes with any COVID-19 fatalities.

The odds of experiencing COVID-19 attributable deaths were higher in homes providing nursing services (OR: 1.8, 95%CI: 1.6 to 2.0); to older people and/or with dementia (OR: 5.5, 95%CI: 4.4 to 6.8); among larger (vs. small) homes (OR: 13.3, 95%CI: 11.5 to 15.4); belonging to a large provider/brand (OR: 1.2, 95%CI: 1.1 to 1.3). There was no significant association with for-profit status of providers.

**Interpretation:** To limit excess mortality, policy should be targeted at care homes to minimise the risk of ingress of disease and limit subsequent transmission. Our findings provide specific characteristic targets for further research on mechanisms and policy priority.

**Funding:** NIHR.

*Summary box:* Evidence before this study
Globally, residents in care homes have experienced disproportionately high morbidity and mortality from COVID-19. Excess mortality incorporates all direct and indirect mortality effects of the pandemic. We searched MEDLINE for published literature, pre-publication databases (medRxiv and Lancet pre-print) and grey literature (ONS and Google) for care homes AND COVID-19 AND mortality, to 31^st^ October 2020. We screened for evidence on excess deaths in care homes in England, and international evidence of the association of COVID-19 deaths and outbreaks with care home characteristics. Official estimates from England and Wales have reported aggregated excess deaths by place of occurrence, but we identified no peer-reviewed excess deaths study in this setting. These aggregates, however, do not account for care home residents dying in other settings (e.g. hospital), nor provide sufficient information to reflect on the impacts of enacted policies over the period, or to inform new policies for future virus waves. Previous peer-reviewed and pre-publication studies have also shown the heterogeneous effects of COVID-19 by care home characteristics in other countries. Particularly important from the current literature appears to be care home size, with larger care homes tending to be associated with more negative outcomes in studies with smaller sample sizes. A study from the Lothian region of Scotland additionally found excess deaths concentrated in a minority of homes that experienced an outbreak. However, a national breakdown of excess deaths by care home characteristics is largely lacking from the current literature in England, with a specific market structure and policy context. Added value of this study
We use nationally representative administrative data from all care homes in England to estimate overall excess deaths and by care home characteristics: setting type (nursing or residential home), client types (offering services for people aged 65+ and/or people with dementia or offering services to children and adults), ownership status (whether not-for-profit - charity/NHS/LA-run homes - or for-profit), whether known to be affiliated to a large provider/brand or independent, and classification according to their registered maximum bed capacity (small, medium and large). We then used multivariable logistic regression to estimate the adjusted odds of a care home experiencing a suspected or confirmed COVID-19 death across these characteristics. We found that only 65% of excess deaths were flagged as officially confirmed/suspected COVID-19 attributed. However, almost all excess deaths occurred in the roughly quarter of care homes that reported at least one suspected/confirmed COVID-19 death. After adjusting for other care home characteristics, larger care homes (vs. small) had the highest odds of experiencing at least one suspected/confirmed COVID-19 death. These findings confirm those from the previous literature, in a unique policy context and with national data. Implications of all the available evidence
The fact that nearly all excess deaths occurred in care homes with at least one COVID-19 attributed death suggests that directly-attributed deaths are very likely to be under-recorded. It also suggests that any indirect mortality effect, of COVID-19 and any enacted policies, were predominantly constrained to those homes experiencing an outbreak. Larger homes are likely to experience higher footfall in general, and so higher probability of contact with an infected individual, which is likely a contributing factor to the association. Furthermore, it might be easier to ensure person-centred protocols in small care homes due to the scale. There is an urgent need for further research to explore the mechanisms in relation to care home characteristics. Also, to empirically test effective interventions, in consideration of additional impacts on quality of life and psychological wellbeing. However, until this is possible, prioritising existing resources, such as testing and PPE equipment, for care homes to prevent ingress of disease is key to preventing large excess mortality.

## 1. Introduction

Globally, residents in care homes have experienced disproportionately high morbidity and mortality from COVID-19. Across Europe, countries adopting more relaxed strategies to tackle the pandemic, such as Sweden, and those adopting severe lockdowns, like Spain and the UK, have both struggled to protect vulnerable persons in care homes.^1,2^ Early international evidence suggests that nearly half of all COVID-19 deaths in five European countries were among care home residents.^2^

Directly attributed COVID-19 deaths do not necessarily capture the full impact on mortality, however.^3^ Death toll for COVID-19 relies on SARS-CoV-2 testing, with tests particularly supply-constrained in early parts of the pandemic. Indirect fatalities due to non-COVID-19-related causes might also have increased. For example, through increased risks of harm from isolation^4^ and possible delayed/cancelled hospital admissions resulting in unintended iatrogenic events and further deaths. Excess deaths, the additional deaths observed in a given period compared to the number usually expected, better capture direct and indirect mortality impacts.

The Office for National Statistics (ONS) in England and Wales have reported aggregated excess deaths by place of occurrence. There was approximately a 79% increase in total deaths in care homes in England and Wales from 2^nd^ March to 12^th^ June compared to 2015-19.^5,6^ These aggregates, however, do not account for care home residents dying in other settings (e.g. hospital), nor provide sufficient information to reflect on the impacts of enacted policies over the period, or to inform new policies for future virus waves.

Likely, not all care homes suffered equally from COVID-19.^7^ In Canada, for-profit status was associated with the number of residents infected and deaths after an outbreak.^8^ Nursing homes with higher nurse-staffing hours per resident were less likely to experience outbreaks across eight US states.^9^ In 179 UK care homes, lower infection rates were found in small homes with high staff-to-resident ratios and low bed occupancy rates.^10^ Large care homes experienced higher rates of infection in Wales,^11^ and in the Lothian region of Scotland excess deaths were concentrated in a minority of care homes with an outbreak.^12^ A national breakdown of excess deaths by care home characteristics is largely lacking from the current literature in England.^6^

The aim of this study is to use nationally representative administrative data from all care homes in England to quantify the excess mortality for residents during the first 23-weeks of the COVID-19 outbreak, and to explore associations with care home characteristics. We then use multivariable logistic regression to estimate the odds of care homes experiencing a suspected or confirmed COVID-19 death across care home types. This knowledge might inform a targeted policy response in future waves.

## 2. Data & Methods

### Institutional context

The English care home market is mostly private and very fragmented, with the type of service provided, client type(s) covered and bed capacity varying systematically by provider-type and the local authorities in which they operate.^13^

There are two main categories of care homes: care homes providing nursing services and those which do not. Care homes which do provide nursing services (nursing homes) cater for people who have complex clinical needs that require regular attention from registered nurses. Care homes which do not offer nursing care (referred to as residential homes) cater for people who often require personal care only, with district nurses and physicians called in when necessary.

Further differences in care homes’ organisational characteristics and operational strategies (for-profit/not-for-profit, independent or belonging to a corporate chain/groups of providers(branded), small/medium/large) might also influence the ability of care homes to put in place effective infection, prevention and control protocols. For example, advanced care planning to ensure patient-centred management, ability to access Personal Protective Equipment (PPE),SARS-CoV-2 testing capacity constraints, as well as staff-to-resident ratios and policies on staff and patient movement across facilities.^14^

Policies adopted in the wider health and care system might have also impacted COVID-19 infections in English care homes. In mid-March, Hospital Trusts discharged medically fit patients to care homes to free capacity.^15^ Mandatory testing prior to discharge was only brought into effect a month later.^16,17^ On March 24^th^ the wider population were ordered not to leave their home except for “essential” reasons,^18^ including visiting care homes, later clarified to only in exceptional circumstances.

### Data

Care home-level daily death notification data sent by registered care home operators in England in the period 1^st^ of January 2017 to 7^th^ August 2020 to the Care Quality Commission (CQC), the independent regulator of health and adult social care in England. All providers must send their notifications to CQC without delay.^19^

The data includes all deaths of care home residents regardless of whether they occurred in care homes or elsewhere (e.g. in hospital).^20^ From 10^th^ April 2020, deaths suspected (based on the statement of care home providers) or confirmed (tested) to be attributable to COVID-19 (COVID-19 deaths) were also identified.^21^ All other deaths are classified as non-COVID-19 deaths.

Death notification data were linked at a care home-level with CQC registers of active care homes in England, providing data on care home characteristics: setting type (nursing or residential home), client types (offering services for people aged 65+ and/or people with dementia or offering services to children and adults), ownership status (whether not-for-profit - charity/NHS/local-authority-run homes - or for-profit), whether known to CQC to be independent or affiliated to a large provider/brand, and their registered maximum bed capacity (coded as small (less than 23 beds), medium (24- 40 beds) and large (41 or more beds)).

Providers enter and leave the market over time. In January 2017, there were 16,481 active care homes, reducing to 15,554 by January 2020. Bed capacity is more stable, with 460,323 beds in January 2017 and 457,347 in January 2020. For the calculation of excess deaths, we use data from the 13,630 care homes which reported at least one death over the study period. 32 (0.17%) care homes were excluded from the analysis due to inability to match with CQC registers. Of all 19,271 care homes reported to be active at some point between 1^st^ of January 2017 and 7^th^ August 2020, 3,747 care homes were no longer active in March 2020, meaning 15,524 care home were used when modelling the odds of experiencing any COVID- 19 fatalities.

To calculate excess deaths, we aggregated daily care home-level deaths to weekly and local authority level. This aimed to reduce the incidence of zeros and the non-constant intra-week variation in death counts (Appendix 1,p.1). Therefore, excess deaths were estimated using aggregated data for 150 local authorities for a period of 188 weeks: 165 weeks (1 January 2017- 3 March 2020) as the pre-COVID-19 period and 23 weeks (4 March 2020 – 7 August 2020) as the post-COVID-19 period, with the start of the COVID-19 period defined by the first week in which one COVID-19 death was reported in England.^22^

### Methods

To calculate excess deaths overall and by care home characteristics, we first used data from the pre-COVID-19 period to estimate expected death trends.^23^ After comparing predictive accuracy with more complex models structures (Appendix 2,p.3), a Poisson regression model (standard log link) was selected, with covariates including a quartic polynomial of week-of-the-year (to account for seasonality) and local authority fixed effects (to account for determinants of deaths that differ across local authorities but do not change over time). Predicted weekly deaths for each local authority were used as the estimated counterfactual in the COVID-19 period (i.e. the deaths that would have occurred in the absence of the pandemic). National excess deaths were computed as the sum of the difference between observed and counterfactual deaths over all local authority-weeks. Excess deaths per hundred bed capacity (excess deaths per bed) were also reported. 95% confidence intervals were constructed by bootstrapping with local authority resampling (50 replications). We also report observed weekly deaths flagged as confirmed or suspected COVID-19 fatalities to show the proportion of excess deaths directly attributed to COVID.

The timing and pattern of excess deaths were likely to vary considerably according to whether a care home has experienced an outbreak or not. As nationwide care home-level data on COVID-19 outbreaks are collated but not publicly available,^24^ we classified care homes according to whether a confirmed/suspected COVID-19 death had been reported in the 23-week COVID-19 period. We used multivariable logistic regression with local-authority fixed effects (controlling for all time-invariant determinants of disease spread at the area level) to estimate unadjusted and adjusted odds of reporting any COVID-19 fatalities on covariates.

All analyses were performed in StataMP v14·2.

## 3. Results

### Descriptive statistics

Table 1 summarises the characteristics of all care homes in England active in March 2020. The minority (4,428, 28.5%) provided nursing services. However, on average nursing homes had more beds than residential homes (50.6 beds versus 20.9) since they are larger (60.2% versus 12.9% with 41+ beds). This explains the more comparable supply of total beds in nursing (223,917) and residential care homes (231,677).

Almost all (93.4%) nursing homes in England provided services to older people and/or people with dementia, as well as the majority (63.1%) of residential homes.

Over a third (37.7%) of care homes were affiliated to a larger branded provider/chain. The proportion of branded care homes was higher (45.8%) for nursing care homes than for residential homes (34.5%). 90.5% of nursing homes were for-profit, with slightly less (83.3%) residential homes.

Overall, approximately 6 in 10 care homes experienced at least one death in the COVID-19 period, with a larger share of nursing homes (89%) reporting fatalities than residential homes (47.9%). 5,641 care homes were active at some point in the study period but did not report fatalities. These were mainly small residential homes, with an average bed capacity of 9.21 (95% CI: 8.95 to 9.49). 27.4% of care homes reported COVID-19 confirmed/suspected fatalities, most in nursing (54.2%) rather than residential homes (16.7%).

**Table 1:** Characteristics of the care homes in England

|  | Overall<br>(Nursing and Residential homes<br>combined) | Nursing<br>homes | Residential<br>homes |
| --- | --- | --- | --- |
| Care homes | 15,524 | 4,428 | 11,096 |
| Average bed capacity | 29.3 | 50.6 | 20.9 |
| Total bed capacity offered | 455,594 | 223,917 | 231,677 |
| Care home size |  |  |  |
| Small homes [0-23 beds] | 49.2% | 12.0% | 64.1% |
| Medium homes [24-40 beds] | 24.4% | 27.8% | 23.0% |
| Large homes [41+ beds] | 26.4% | 60.2% | 12.9% |
| Service type |  |  |  |
| Providing services to Older People and/or people with<br>dementia | 71.7% | 93.4% | 63.1% |
| Providing non-dementia services to children and/or<br>adults only | 28.3% | 6.6% | 36.9% |
| Provider type |  |  |  |
| Branded (chain ownership) | 37.7% | 45.8% | 34.5% |
| Non-branded (independent ownership) | 62.3% | 54.2% | 65.5% |
| Legal status |  |  |  |
| For-profit | 85.3% | 90.5% | 83.3% |
| Not-for-profit | 14.7% | 9.5% | 16.7% |
| % care homes reporting any death in the COVID-19<br>period | 59.7% | 89.0% | 47.9% |
| % care homes reporting confirmed/suspected COVID-19<br>fatalities | 27.4% | 54.2% | 16.7% |

Notes: Own elaboration on CQC data on care homes reported to be active in March 2020.

### Observed, Expected and Excess deaths

Prior to the COVID-19 period, predicted deaths tracked observed deaths relatively closely (Appendix 3, p.9). During the COVID-19 period, observed deaths were considerably higher than predicted from historical trends (Figure 1). Most excess deaths occurred during the 10 weeks between 25^th^ March and 2^nd^ June. At the end of the study period, excess deaths were lower than predicted, especially in nursing homes. Overall, 64.7% of calculated excess deaths (95% CI: 56.4% to 76.0%) were reported to be attributable to confirmed/suspected COVID-19, with this proportion increasing over time.

**Figure 1:**
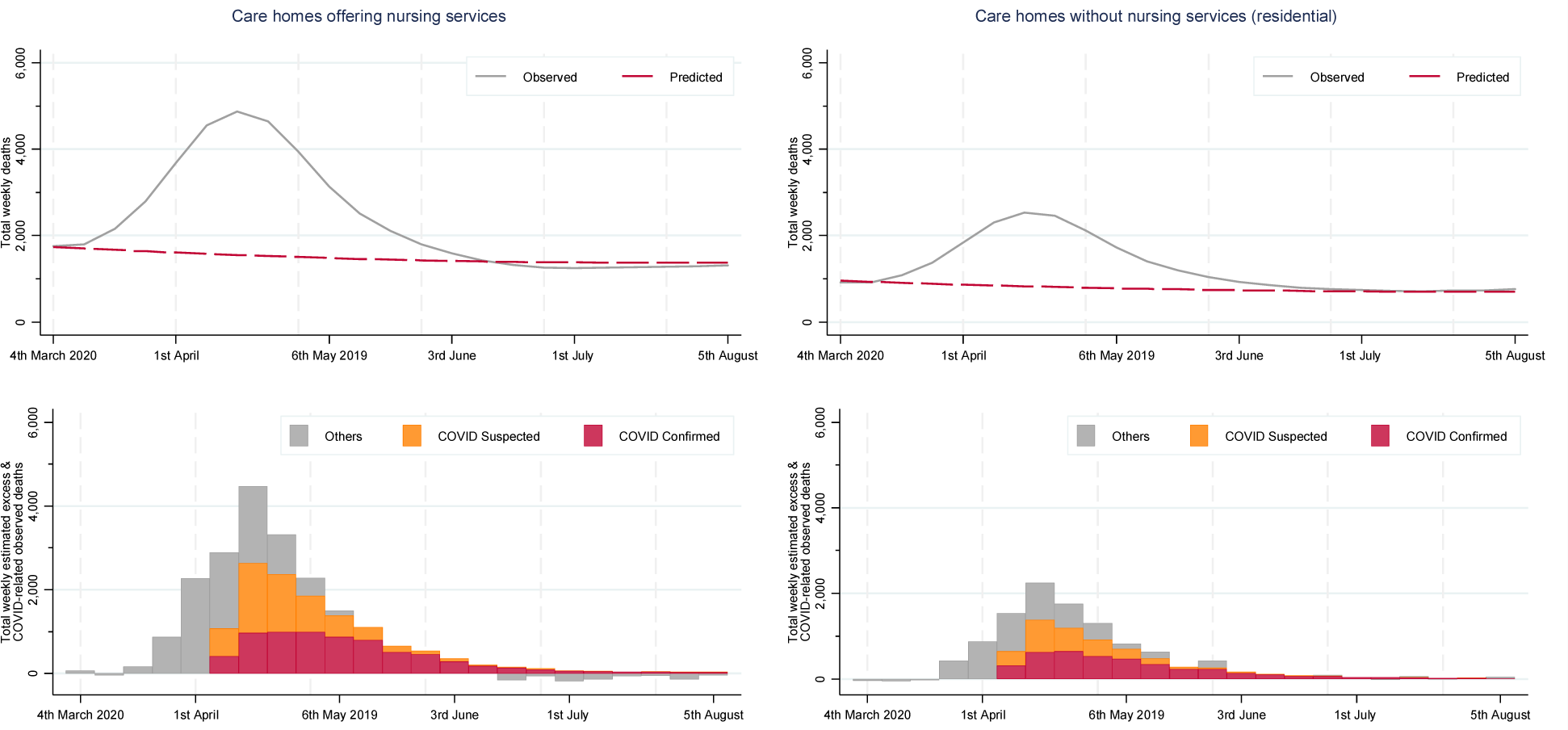
Predicted versus observed deaths, and estimated excess deaths by care home setting type in the first 23 weeks of the COVID-19 pandemic in England Notes: smoothed weekly reported death counts (observed) and predicted deaths before and after week 166 obtained from local linear regressions.

There were 29,542 (95% CI: 25,176 to 33,908) excess deaths in all care homes over the COVID-19 period (Table 2), equivalent to excess deaths per bed of 6.3% (95% CI: 5.4% to 7.2%). Robustness checks (Appendix 2,p.3) showed consistent excess death estimates across different modelling approaches (range: 29,542 to 29,711).

Excess deaths were higher in nursing (18,891, 95% CI: 15,956 to 21,826) compared to residential homes (10,651, 95% CI: 8,914 to 12,388), with almost double excess deaths per bed (8.4% versus 4.6%).

Excess deaths were significantly higher in facilities that provided services to older people and people with dementia, mainly for nursing homes, with excess deaths per bed of 8.6% (95% CI: 7.3% to 9.9%).

For-profit and not-for-profit nursing homes had comparable excess deaths per bed, but the rate was nearly double in not-for-profit residential homes (7.7% versus 4.1%).

Branded homes experienced higher excess deaths per bed than independent care homes (7.2% versus 5.8%). This difference largely occurred among residential homes (5.6% versus 4%).

Larger care homes also had higher excess deaths per bed (8.6% versus 2.2% in small homes).

The starkest difference in excess deaths was between care homes that experienced and did not experience at least one suspected/confirmed COVID-19 death, with the former responsible for almost all excess deaths (29,429, 95% CI: 25,047 to 33,810). For graphical inspection see Appendix 4,p.12. For homes experiencing COVID-19 attributable fatalities, excess deaths per bed were comparable across settings (13.8% nursing; 13.0% residential). Estimated excess deaths for nursing homes not reporting COVID-19 deaths were negative (−831, 95% CI: -1,291 to -371), corresponding to an estimated excess deaths per bed of -1% (95% CI: -1.6% to - 0.5%). On the other hand, there were 943 excess deaths (95% CI: 503 to 1,383) in residential homes not reporting COVID-19 deaths, corresponding to an excess deaths per bed of 0.6% (95% CI: 0.3% to 0.9%).

**Table 2:** Excess deaths (panel a) and excess deaths per bed (panel b) by care home type (in the first 24 weeks with COVID-19)

a) Total excess deaths*
|  | Overall (Nursing and Residential homes combined) | Nursing homes | Residential homes |
| --- | --- | --- | --- |
| Overall excess death | 29,542 [25,176;33,908] | 18,891 [15,956;21,826] | 10,651 [8,914;12,388] |
| % excess deaths attributable to COVID-19 | 64.7 [56.41;75.98] | 67.1 [58.11;79.49] | 60.5 [52.03;72.30] |
| Reporting COVID-19 deaths | 29,429 [25,047;33,810] | 19,722 [16,665;22,778] | 9,707 [8,011;11,403] |
| Not reported | 112 [-457;682] | -831 [-1,291;-371] | 943 [503;1,383] |
| Providing services to older people / with dementia | 28,958 [24,673;33,242] | 18,786 [15,858;21,714] | 10,171 [8,470;11,873] |
| Providing non-dementia services to children and/or adults only | 584 [380;789] | 105 [44;166] | 479 [292;666] |
| For-Profit care homes | 25,468 [21,685;29,251] | 17,335 [14,782;19,888] | 8,133 [6,592;9,674] |
| Not-for-profit care homes | 4,074 [3,081;5,067] | 1,556 [858;2,255] | 2,517 [2,005;3,030] |
| Branded care homes | 14,671 [12,053;17,288] | 9,776 [8,017;11,534] | 4,895 [3,829;5,962] |
| Non-branded care homes | 14,871 [12,702;17,041] | 9,116 [7,540;10,692] | 5,755 [4,864;6,647] |
| Small homes [0-23 beds] | 1,752 [1,410;2,095] | 195 [55;335] | 1,558 [1,231;1,885] |
| Medium homes [24-40 beds] | 5,751 [4,791;6,711] | 2,480 [1,869;3,092] | 3,270 [2,770;3,771] |
| Large homes [41+ beds] | 22,039 [18,564;25,513] | 16,216 [13,704;18,729] | 5,823 [4,524;7,122] |

*Panel b) total excess deaths per bed (in %, adjusted for bed capacity as reported in March 2020)*
|  | Overall (Nursing and Residential homes combined) | Nursing homes | Residential homes |
| --- | --- | --- | --- |
| Overall excess | 6.5 [5.5;7.4] | 8.4 [7.1;9.7] | 4.6 [3.8;5.3] |
| Overall excess attributable to COVID-19 | 4.2 [3.1;5.7] | 5.7 [4.1;7.7] | 2.8 [2.0;3.9] |
| Reporting COVID-19 deaths | 13.5 [11.5;15.5] | 13.8 [11.7;15.9] | 13.0 [10.7;15.2] |
| Not reported | 0.0 [-0.2;0.3] | -1.0 [-1.6;-0.5] | 0.6 [0.3;0.9] |
| Providing services to older people / with dementia | 7.0 [5.9;8.0] | 8.6 [7.3;9.9] | 5.1 [4.3;6.0] |
| Providing non-dementia services to children and/or adults only | 1.5 [1.0;2.0] | 1.9 [0.8;3.0] | 1.4 [0.9;2.0] |
| For-Profit care homes | 6.3 [5.4;7.2] | 8.4 [7.2;9.7] | 4.1 [3.3;4.9] |
| Not-for-profit care homes | 8.0 [6.1;10.0] | 8.7 [4.8;12.6] | 7.7 [6.1;9.2] |
| Branded care homes | 7.2 [6.0;8.5] | 8.4 [6.9;10.0] | 5.6 [4.4;6.9] |
| Non-branded care homes | 5.8 [5.0;6.7] | 8.4 [7.0;9.9] | 4.0 [3.4;4.6] |
| Small homes [0-23 beds] | 2.2 [1.8;2.7] | 2.6 [0.7;4.5] | 2.2 [1.7;2.6] |
| Medium homes [24-40 beds] | 4.7 [4.0;5.5] | 6.0 [4.6;7.5] | 4.1 [3.5;4.7] |
| Large homes [41+ beds] | 8.6 [7.3;10.0] | 9.2 [7.8;10.7] | 7.3 [5.6;8.9] |

### Care home characteristics associated with odds of one COVID-19 death

After adjusting for other care home characteristics, nursing homes had a statistically significant higher odds of experiencing COVID-19 confirmed/suspected deaths than residential homes (OR: 1.8, 95% CI: 1.6 to 2.0) (Table 3). Care homes offering services to older people and/or people with dementia were particularly exposed (OR: 5.5, 95% CI: 4.4 to 6.8). Branded care homes experienced significantly higher odds of one COVID-19-related death compared to independent homes (OR: 1.2, 95% CI: 1.1 to 1.3), although not-for-Profit and for-profit homes experienced roughly equal odds of one COVID-19 death (overall OR: 1.0, 95% CI: 0.8 to 1.1). Compared to small care homes, medium-sized facilities experienced higher odds of COVID-19- related deaths (OR: 5.2, 95% CI: 4.5 to 6.0), with large-sized homes experienced even greater odds (OR: 13.3, 95% CI: 11.5 to 15.4). Results were robust to controlling for deprivation and urbanicity of the care home location, and to restricting the sample to care homes providing services to older people/with dementia (Appendix 5, p.14).

**Table 3:** The odds ratios (and 95%CI) of experiencing COVID-19 confirmed/suspected deaths in the English care homes

|  | % of care homes in each category reporting COVID-19 deaths | Unadjusted OR (95%CI) |  |  | Multivariable adjusted OR (95%CI) |  |  |
| --- | --- | --- | --- | --- | --- | --- | --- |
|  |  | OR | 95% CI | p-value | OR | 95% CI | p-value |
| Overall (Nursing and Residential care homes combined) |  |  |  |  |  |  |  |
| Providing nursing services | 20.0 | 5.93 | (5.47-6.42) | <0.001 | 1.81 | (1.64-1.99) | <0.001 |
| Providing residential services only | 14.9 | 1 | (reference) |  | 1 | (reference) |  |
| Providing services to older people/with dementia | 17.0 | 26.87 | (21.95-32.88) | <0.001 | 5.45 | (4.36-6.81) | <0.001 |
| Providing non-dementia services to children and/or adults only | 19.2 | 1 | (reference) |  | 1 | (reference) |  |
| Not-for-profit care home | 21.2 | 0.54 | (0.48-0.61) | <0.001 | 0.96 | (0.83-1.11) | 0.605 |
| For-profit care homes | 16.6 | 1 | (reference) |  | 1 | (reference) |  |
| Branded care homes | 21.7 | 1.6 | (1.48-1.72) | <0.001 | 1.21 | (1.1-1.34) | <0.001 |
| Non-branded care homes | 14.4 | 1 | (reference) |  | 1 | (reference) |  |
| Small homes [0-23 beds] | 11.4 | 0.05 | (0.04-0.05) | <0.001 | 1 | (reference) |  |
| Medium homes [24-40 beds] | 14.7 | 1.55 | (1.43-1.69) | <0.001 | 5.2 | (4.52-5.98) | <0.001 |
| Large homes [41+ beds] | 21.9 | 10.34 | (9.49-11.27) | <0.001 | 13.28 | (11.46-15.39) | <0.001 |
| Nursing care homes |  |  |  |  |  |  |  |
| Providing services to older people/with dementia | 20.0 | 9.99 | (6.93-14.42) | <0.001 | 2.98 | (1.98-4.49) | <0.001 |
| Providing non-dementia services to children and/or adults only | 22.7 | 1 | (reference) |  | 1 | (reference) |  |
| Not-for-profit care home | 24.8 | 0.59 | (0.47-0.73) | <0.001 | 0.91 | (0.71-1.17) | 0.48 |
| For-profit care homes | 19.6 | 1 | (reference) |  | 1 | (reference) |  |
| Branded care homes | 23.1 | 1.61 | (1.42-1.83) | <0.001 | 1.26 | (1.1-1.45) | 0.001 |
| Non-branded care homes | 17.4 | 1 | (reference) |  | 1 | (reference) |  |
| Small homes [0-23 beds] | 14.5 | 0.09 | (0.07-0.11) | <0.001 | 1 | (reference) |  |
| Medium homes [24-40 beds] | 16.3 | 0.56 | (0.48-0.64) | <0.001 | 4.39 | (3.21-5.99) | <0.001 |
| Large homes [41+ beds] | 22.1 | 4.1 | (3.58-4.69) | <0.001 | 10.88 | (8.03-14.74) | <0.001 |
| Residential care homes |  |  |  |  |  |  |  |
| Providing services to older people/with dementia | 14.7 | 21.8 | (16.97-28) | <0.001 | 6.57 | (5-8.63) | <0.001 |
| Providing non-dementia services to children and/or adults only | 18.2 | 1 | (reference) |  | 1 | (reference) |  |
| Not-for-profit care home | 19.5 | 0.73 | (0.63-0.85) | <0.001 | 0.99 | (0.83-1.19) | 0.943 |
| For-profit care homes | 14.3 | 1 | (reference) |  | 1 | (reference) |  |
| Branded care homes | 20.2 | 1.31 | (1.17-1.46) | <0.001 | 1.16 | (1.02-1.33) | 0.028 |
| Non-branded care homes | 12.8 | 1 | (reference) |  | 1 | (reference) |  |
| Small homes [0-23 beds] | 10.9 | 0.07 | (0.06-0.08) | <0.001 | 1 | (reference) |  |
| Medium homes [24-40 beds] | 14.0 | 2.86 | (2.56-3.2) | <0.001 | 5.13 | (4.37-6.02) | <0.001 |
| Large homes [41+ beds] | 21.5 | 9.79 | (8.6-11.15) | <0.001 | 13.48 | (11.28-16.11) | <0.001 |

Notes: 15,524 Care homes in England (4,428 nursing and 11,096 residential) reported to be active in March 2020 to CQC. Appendix 5 reports estimates from sensitivity analysis.

## 4. Discussion

### 4.1. Principal Findings

Using provider-level administrative data on all care homes in England, we estimated that there were over 29,500 excess deaths of care home residents during the first 23-weeks of COVID-19, equivalent to 6.5% of all care home beds. Almost 65% of the excess deaths were reported to be directly attributable (confirmed/suspected) to COVID-19. Our analysis shows that almost all excess deaths were recorded in the quarter of care homes which reported COVID-19 fatalities. This highlights that: (i) non-COVID-19 attributed excess deaths were likely to be directly due to COVID-19; and/or (ii) any indirect negative effects of COVID-19 and enacted policies on mortality were predominantly constrained to those homes experiencing an outbreak. Non-COVID-19 attributed deaths being reported mainly during the early stages of the pandemic, when CQC recording of COVID-19 death was missing (before 10^th^ April), guidance focused on a narrower set of symptoms and there was a shortage of testing, providing support for the former hypothesis.

Excess deaths were mainly concentrated among large and branded homes that provide services to older people and people with dementia. Adjusted care home level analysis confirmed these findings.

### 4.2. Strengths & Limitations

To our knowledge, this is the first independent analysis that uses national administrative records from all care homes in England to estimate the impact of COVID-19. We find comparable total deaths to official estimates,^5^ adding stratifications of excess deaths by key care home characteristics and multivariable analysis to add a more nuanced understanding of these deaths. Local-authority fixed effects were used to account for time-invariant measured and unmeasured determinants and confounders that differ across local authority.

Our study also has limitations. Firstly, we can observe the counts of COVID-19 attributed fatalities across care homes but not whether non-fatal COVID-19 cases occurred. This case data is not available, though serological and whole genome sequencing studies give insights to this.^24^ The attribution of COVID-19-related deaths is based on statements from providers to the CQC starting from 10^th^ April 2020 and not always testing-confirmed or reflected in the death certificate. COVID- 19 attributable deaths occurred before the 10^th^ April would have been miscoded. The reported lower rates of testing could lead to some relevant deaths not having COVID-19 listed as a contributory factor, leading to apparently higher non-COVID-19 excess deaths.^5,10,21^

No data was available on occupancy rates at care home level. We instead used maximum bed capacity as reported in March 2020, assuming full occupancy. In the UK, occupancy rates were estimated to be on average 90% in nursing homes and 91% in residential homes.^13^ It is very likely that occupancy rates declined during the COVID-19 period. However, assuming an arbitrary lower occupancy would increase excess mortality rates only proportionally, unless further breakdowns by time and care home types became available.

Measures of staffing and working conditions, residents’ case mix and their socio- demographic status, and individual care home shortages of equipment would have been relevant for this analysis but there is no national care-home level data available. Our analysis is instead based on providers’ characteristics as reported to CQC.

We did not account for exposure and incidence of COVID-19 in the local area where each care home is located, or local policy responses to the pandemic, which changed over time. Wider community testing was negligible in the early parts of our analysis period,^25^ and likely differed by local authority capacity which would bias results if included. Good quality data on local policy responses was also unavailable.

Finally, as the number of deaths in the absence of the outbreak cannot be observed but only predicted, there is the potential that market dynamics and prediction errors could influence excess deaths estimates. However, we estimated small prediction errors in the pre-COVID-19 period relative to the size of excess deaths in the COVID period. Excess death estimates were also robust to different modelling approaches.

### 4.3. Study in context

By comparing observed deaths against averages over historical 5-year period, the ONS estimated 25,876 excess deaths in English care homes up to August 8^th^.^5^ Our estimates exceed this slightly. In addition to differences in methods, this is likely due to our data including deaths of care home residents occurring outside of a care home setting (e.g. in hospital).

Consistent with previous studies, we find that excess deaths occur overwhelmingly in the minority of care homes that experience COVID-19 fatalities.^12^ This might suggest higher proportions of COVID-19-related excess deaths than reported,^26^ and that some death are potentially avoidable if initial care home outbreaks had been prevented.

However, our results suggest that other care homes characteristics, relating to type of residents, staffing, ownership and size, are also important.

Care homes providing services to older people/with dementia suffered most deaths. This is unsurprising given the increased risk of contracting SARS-CoV-2 (difficulties complying with physical distancing, masking and hand hygiene) and increased risk of morbidity/mortality (comorbid illnesses), frailty and age. However, for care homes serving this group, there were smaller odds of COVID-19-related deaths in nursing compared with residential care homes. This might suggest a protective effect of the presence of staff with nursing backgrounds and infection, prevention and control (IPC) training, as found in other settings.^9^

Overall, though, nursing homes had the most excess deaths and odds of COVID-19 confirmed/suspected deaths. This is likely due to these homes containing residents at high risk of contracting and dying of SARS-CoV-2, increased frailty and higher prevalence of co-morbidities, and therefore a greater likelihood of being in contact with other healthcare settings and practitioners.^27^

In line with the existing literature, we found that large care homes are more likely to experience negative outcomes.^10-12^ A likely contributor is that larger homes have a higher footfall altogether, of staff, healthcare workers, residents flowing in and out of hospitals, and visitors in non-pandemic times. This increases their chances of exposure to an infected individual, particularly in the absence of rigorous testing. Furthermore, it might be easier to ensure patient-centred management protocols in small care homes where policies around staff and patients contacts are set for smaller scales.^28^

We find no significant differences between for-profit and non-for-profit providers, although for-profit providers experienced the most excess deaths because they account for the majority of the market. A Canadian study showed for-profit status was not associated with the odds of an outbreak, although it was associated with the extent of an outbreak (number of cases and deaths).^8^ However, we find that branded care homes had greater odds of COVID-19 confirmed/suspected deaths and rates of excess deaths. Branded homes could have policies around staff and patient movement across facilities that could aid the spread of infection.^7^

### 4.4. Policy implications & future research

Specialist initiatives are needed for patients/staff/visitors to minimise the risk of initial infection in care homes. What prevention policies are optimal (e.g. PCR testing, staff cohorting, visitor restrictions, hospital discharge policies, limiting visiting professionals, tracing staff etc.)^29^ requires further research and dialog with operators and public health experts. Their efficacy depends upon the care home setting in which they are implemented and the behavioural responses of residents and staff. Critically, any benefits from such policies would need to be weighed against costs and potential adverse outcomes, such as reduced quality-of-life or psychological well-being.^29,30^

There is an urgent need for further research to explore the mechanisms hypothesized above in more detail. Evaluations of alternative interventions are required. However, our results suggest that until this is possible, prioritising existing resources, such as testing and PPE equipment, to prevent initial infections in care homes is key to preventing large excess mortality.

## Data Availability

Care home-level daily death notification data provided by the Care Quality Commission (CQC) and linked with CQC registers of active care homes in England. Neither the funders nor the collectors/curators/delivers of the data bears any responsibility for the analyses or interpretations presented here. Data can be requested to the CQC. The statistical code is available from the corresponding author. 

## Contributors

MM did the econometric analysis, and led the design and the writing of the paper. JS and AJT contributed to the design, the econometric analysis, interpretation of findings and the writing of the paper. IA oversaw the econometric analysis, supported the data acquisition, contributed to the design, interpretation of findings and the writing of the paper. EK advised on the design and contributed to the writing of the paper. AJT and IH verified the underlying data. The authors are grateful to the DH&SC, CQC and the members of the Care Home Working Group for their suggestions in all phases of the research.

## Data Availability

Care home-level daily death notification data provided by the Care Quality Commission (CQC) and linked with CQC registers of active care homes in England. Data Neither the funders nor the collectors/curators/delivers of the data bears any responsibility for the analyses or interpretations presented here. Data can be requested to the CQC. The statistical code is available from the corresponding author.

