## Appendices for "Excess mortality for care home residents during the first 23 weeks of the COVID-19 pandemic in England: a national cohort study"

**Appendix 1: Construction of the analysis sample**

Daily reported death counts follow a strong idiosyncratic intra-week pattern, with low fatalities reported over the weekend/public holidays/long weekends and a spike reported on the next day (See Figure A1-1).^[1]^

***Figure A1-1:*** *Average death toll by day of the week in England*

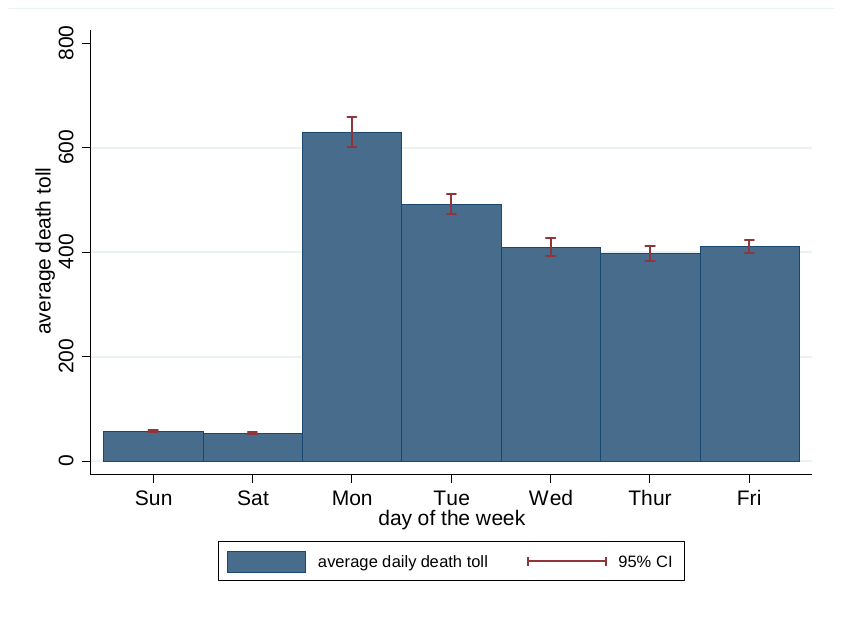

*Notes:* Averages computed over the pre-COVID-19 study period 1^st^ January 2017 to 4^th^ March 2020

To reduce the incidence of zeros and of non-constant intra-week variation, we aggregated daily deaths to weekly level, with January 1^st^ as the start of the first week of a year composed of precisely 52 weeks. Thus weeks 52, 104 and 156 of the study period are 8 days, while all the remaining weeks are usual 7 days. This ensures that each week of a given year overlaps precisely with an equivalent week in other years,^[2]^ with the exception of week 165 of the study period (the week containing February 29^th^), 2020 being a leap year.

Weekly reported death counts overall and by stratum of interest were further aggregated at the level of local authority to i) limit the effect of confounding from supply-side care service provision (e.g. regarding the supply of care beds, and quality and prices of care home provision) on observed mortality;^[3]^ and ii) to reduce the predominance of zeros in weekly death counts. We have excluded data from two local authorities: City of London (no care homes) and Isle of Scilly (one care home providing residential services only).

Our analysed data therefore comprises aggregated data for 150 local authorities (LAs) for a full time horizon of 188 weeks: 165 weeks (1 January 2017- 3 March 2020) as the pre-COVID-19 period and the following 23 weeks (first week with a COVID-19 death^25^ 4 March 2020 – 7 August 2020) as the post-COVID-19 period. Figure A1-2 reports variation in daily death toll across local authorities, unadjusted for bed capacity. This suggest a statistical framework with fixed local-authority effects and the analysis of excess deaths relative to bed capacity.

***Figure A1-2:*** *Variation in the weekly death toll across local authorities*
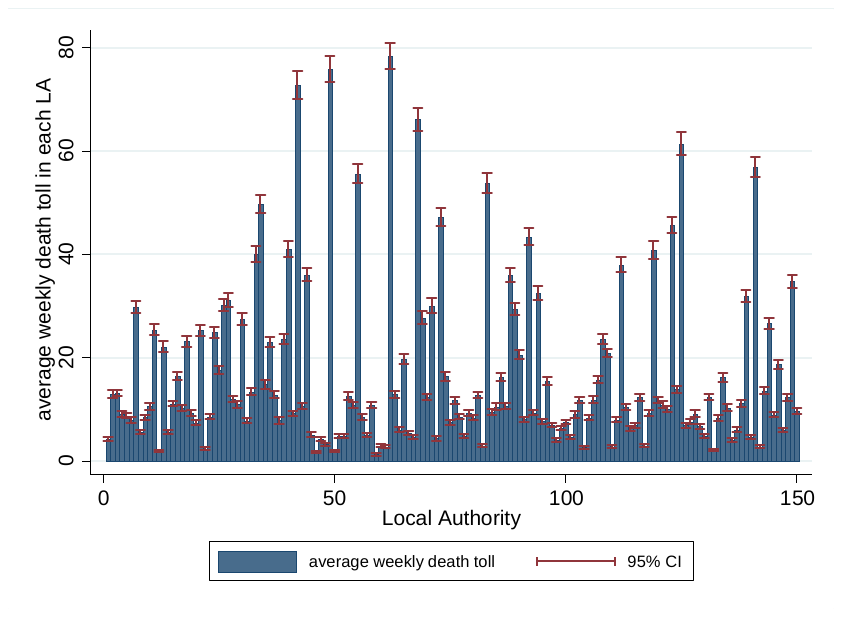

*Notes:* Averages computed over the pre-COVID-19 study period 1^st^ January 2017 to 4^th^ March 2020.

**Appendix 2: Model selection and robustness to alternative specifications of seasonality**

The optimal econometric specification used to estimate counterfactual deaths in the COVID-19 period was chosen based on split-sample validation. This involved separating the pre-COVID-19 period into an estimation sample (1^st^ January 2017 to 4^th^ March 2019) and a hold-out sample (5^th^ March 2019 to 12^th^ August 2019), with the latter chosen to most closely reflect a comparable COVID-19 period in the year prior to COVID-19. Estimates obtained in the estimation sample were used to obtain out-of-sample predictions for the hold-out sample to assess the accuracy of our predictions. Performance of each model was assessed by computing the mean error (ME) and root mean squared error (RMSE) in the hold-out sample, with the optimal model chosen as the model which minimised the RMSE.

We tested the performance of a series of specifications differing in how seasonality and time trends were accounted for. We considered the following functional forms:

1. A quartic (fourth degree) polynomial of week-of-the-year with no time trend (assuming stationarity)
2. A quartic (fourth degree) polynomial of week-of-the-year with a linear time (week) trend
3. A quartic (fourth degree) time (week) polynomial
4. Week-of-the-year fixed effects with a linear time (week) trend

Here, week-of-the-year takes one of 52 values, with week one representing the first week in January, and week 52 representing the final week in December. Given the estimation sample runs from 1st January 2017 to 4th March 2019, there are three data points for weeks 1 to 9 for each local authority, and two data points for the remaining weeks. For the specification including week-of-the-year fixed effects, the week commencing 1st January is used as the reference category, with 51 binary indicators for each of the remaining weeks-of-the-year included.

The time (week) variable represents a week counter running from a value of one (representing the week commencing 1st January 2017) to a value of 113 (week commencing 25th February 2019), which is the final week of the estimation sample.

For specifications featuring quartic polynomials, we include linear, squared, cubic, and “to the power 4” terms for either week (specification 3) or week-of-the-year (specifications 1 and 2).

Eights models were explored in total. The first four include functional forms 1. to 4., with the addition of local authority fixed effects (allowing a constant shift in the levels of predicted deaths across local authorities). We also tested the assumption of constant seasonality and time trends across local authorities using specifications which include local authority-specific seasonality and time trends. The second four models therefore interacted functional forms 1. to 4. with local authority fixed effects to allow for local authority-specific seasonality and time trends.

We also explored the performance of a further eight models, including local authority-level average bed occupancy as an exposure term.

The validation exercise was run separately for deaths in all care homes and separately by setting type (residential and nursing care homes).

Model performance for predicting deaths in the hold-out sample for all care homes combined are shown in Table A2.1. These show that the specification with a stationary quartic week-of-the-year polynomial, local authority fixed effects and no exposure term had the lowest RMSE. This specification also minimised the RMSEs when predicting deaths in residential care homes (Table A2.2) and nursing care homes (Table A2.3). This specification performed well on the ME, indicating that predicted and observed deaths were close to identical on average across the hold-out sample. This specification was therefore used for estimating excess deaths reported in the paper.

In equation from, this model can be described as follows:

$$f\left( D_{l,t} \right)=\frac{e^{-exp(\theta_{l,t})}e^{\theta_{l,t}D_{l,t}}}{D_{l,t}!}$$

$$\theta_{l,t}=\beta_{1}woy+\beta_{2}{woy}^{2}+\beta_{3}{woy}^{3}+\beta_{4}{woy}^{4}+\alpha_{l}$$

Where $D_{l,t}$ represents the number of care home deaths in local authority $l=1, \ldots, 150)$ in week $t (=1, \ldots, 165)$, $\alpha_{l}$ represents the set of local authority fixed effects, and $woy, {woy}^{2}, {woy}^{3}$, and ${woy}^{4}$ represent the quartic polynomial of week-of-the-year.

***Table A2-1:*** *Results of the validation exercise for deaths in all care homes*

| **Model** | | |  | **Model performance**** | | | |
| --- | --- | --- | --- | --- | --- | --- | --- |
| **Estimation model** | **Local authority-specific seasonality and time trend?** | **Specification*** |  | **ME** | **RMSE** | **Rank (ME)** | **Rank (RMSE)** |
| **OLS** | No | 1 |  | 0.15 | 5.25 | 1 | 13 |
|  |  | 2 |  | 0.61 | 5.28 | 14 | 14 |
|  |  | 3 |  | -29.66 | 34.50 | 20 | 19 |
|  |  | 4 |  | 0.65 | 5.37 | 18 | 15 |
|  | Yes | 1 |  | 0.15 | 5.03 | 2 | 6 |
|  |  | 2 |  | 0.61 | 5.19 | 15 | 12 |
|  |  | 3 |  | -29.66 | 49.30 | 19 | 20 |
|  |  | 4 |  | 0.65 | 6.25 | 17 | 18 |
| **Poisson** | No | 1 |  | 0.16 | 4.95 | 3 | 1 |
|  |  | 2 |  | 0.55 | 5.00 | 9 | 3 |
|  |  | 3 |  | -112.88 | 238.34 | 21 | 21 |
|  |  | 4 |  | 0.57 | 5.16 | 11 | 10 |
|  | Yes | 1 |  | 0.16 | 5.02 | 4 | 4 |
|  |  | 2 |  | 0.52 | 5.12 | 7 | 8 |
|  |  | 3 |  | -139.27 | 345.63 | 23 | 23 |
|  |  | 4 |  | 0.53 | 6.16 | 8 | 16 |
| **Poisson, exposure** | No | 1 |  | 0.24 | 4.98 | 5 | 2 |
|  |  | 2 |  | 0.59 | 5.02 | 13 | 5 |
|  |  | 3 |  | -114.90 | 243.92 | 22 | 22 |
|  |  | 4 |  | 0.61 | 5.18 | 16 | 11 |
|  | Yes | 1 |  | 0.25 | 5.04 | 6 | 7 |
|  |  | 2 |  | 0.56 | 5.14 | 10 | 9 |
|  |  | 3 |  | -143.31 | 359.90 | 24 | 24 |
|  |  | 4 |  | 0.58 | 6.17 | 12 | 17 |
| *Specification 1 includes a quartic (fourth degree) polynomial of week-of-the-year; specification 2 includes a quartic polynomial of week-of-the-year and a linear time (week) trend; specification 3 includes a quartic time (week) polynomial; and specification 4 includes week-of-the-year fixed effects with a linear time (week) trend.  **All models are estimated using data from 1^st^ January 2017 to 4^th^ March 2019, and model performance is assessed in the period 5^th^ March 2019 to 12^th^ August 2019. | | | | | | | |

***Table A2-2:*** *Results of the validation exercise for deaths in nursing care homes*

| **Model** | | |  | **Model performance**** | | | |
| --- | --- | --- | --- | --- | --- | --- | --- |
| **Estimation model** | **Local authority-specific seasonality and time trend?** | **Specification*** |  | **ME** | **RMSE** | **Rank (ME)** | **Rank (RMSE)** |
| **OLS** | No | 1 |  | 0.16 | 4.01 | 2 | 9 |
|  |  | 2 |  | 0.38 | 4.02 | 12 | 10 |
|  |  | 3 |  | -17.67 | 20.65 | 20 | 19 |
|  |  | 4 |  | 0.40 | 4.08 | 15 | 15 |
|  | Yes | 1 |  | 0.16 | 3.97 | 1 | 6 |
|  |  | 2 |  | 0.38 | 4.05 | 11 | 13 |
|  |  | 3 |  | -17.67 | 29.74 | 19 | 20 |
|  |  | 4 |  | 0.40 | 4.88 | 16 | 18 |
| **Poisson** | No | 1 |  | 0.16 | 3.90 | 3 | 1 |
|  |  | 2 |  | 0.36 | 3.92 | 9 | 2 |
|  |  | 3 |  | -58.17 | 118.08 | 21 | 21 |
|  |  | 4 |  | 0.37 | 4.04 | 10 | 12 |
|  | Yes | 1 |  | 0.16 | 3.96 | 4 | 5 |
|  |  | 2 |  | 0.32 | 4.00 | 8 | 8 |
|  |  | 3 |  | -79.88 | 220.36 | 23 | 23 |
|  |  | 4 |  | 0.32 | 4.82 | 7 | 16 |
| **Poisson, exposure** | No | 1 |  | 0.19 | 3.93 | 6 | 3 |
|  |  | 2 |  | 0.43 | 3.95 | 17 | 4 |
|  |  | 3 |  | -59.54 | 122.12 | 22 | 22 |
|  |  | 4 |  | 0.44 | 4.06 | 18 | 14 |
|  | Yes | 1 |  | 0.19 | 3.99 | 5 | 7 |
|  |  | 2 |  | 0.40 | 4.03 | 13 | 11 |
|  |  | 3 |  | -87.16 | 273.12 | 24 | 24 |
|  |  | 4 |  | 0.40 | 4.84 | 14 | 17 |
| *Specification 1 includes a quartic (fourth degree) polynomial of week-of-the-year; specification 2 includes a quartic polynomial of week-of-the-year and a linear time (week) trend; specification 3 includes a quartic time (week) polynomial; and specification 4 includes week-of-the-year fixed effects with a linear time (week) trend.  **All models are estimated using data from 1^st^ January 2017 to 4^th^ March 2019, and model performance is assessed in the period 5^th^ March 2019 to 12^th^ August 2019. | | | | | | | |

***Table A2-3:*** *Results of the validation exercise for deaths in residential care homes*

| **Model** | | |  | **Model performance**** | | | |
| --- | --- | --- | --- | --- | --- | --- | --- |
| **Estimation model** | **Local authority-specific seasonality and time trend?** | **Specification*** |  | **ME** | **RMSE** | **Rank (ME)** | **Rank (RMSE)** |
| **OLS** | No | 1 |  | -0.01 | 2.85 | 4 | 13 |
|  |  | 2 |  | 0.23 | 2.86 | 15 | 14 |
|  |  | 3 |  | -11.99 | 14.13 | 20 | 19 |
|  |  | 4 |  | 0.25 | 2.88 | 18 | 15 |
|  | Yes | 1 |  | -0.01 | 2.73 | 3 | 5 |
|  |  | 2 |  | 0.23 | 2.84 | 16 | 12 |
|  |  | 3 |  | -11.99 | 21.75 | 19 | 20 |
|  |  | 4 |  | 0.25 | 3.38 | 17 | 18 |
| **Poisson** | No | 1 |  | 0.00 | 2.71 | 2 | 1 |
|  |  | 2 |  | 0.19 | 2.73 | 12 | 3 |
|  |  | 3 |  | -60.69 | 147.25 | 21 | 21 |
|  |  | 4 |  | 0.20 | 2.77 | 14 | 8 |
|  | Yes | 1 |  | 0.00 | 2.73 | 1 | 4 |
|  |  | 2 |  | 0.16 | 2.79 | 8 | 10 |
|  |  | 3 |  | N/A | N/A | 23 | 23 |
|  |  | 4 |  | 0.17 | 3.32 | 10 | 16 |
| **Poisson, exposure** | No | 1 |  | 0.02 | 2.72 | 6 | 2 |
|  |  | 2 |  | 0.18 | 2.73 | 11 | 6 |
|  |  | 3 |  | -61.11 | 148.42 | 22 | 22 |
|  |  | 4 |  | 0.20 | 2.78 | 13 | 9 |
|  | Yes | 1 |  | 0.02 | 2.74 | 5 | 7 |
|  |  | 2 |  | 0.16 | 2.80 | 7 | 11 |
|  |  | 3 |  | N/A | N/A | 24 | 24 |
|  |  | 4 |  | 0.17 | 3.33 | 9 | 17 |
| *Specification 1 includes a quartic (fourth degree) polynomial of week-of-the-year; specification 2 includes a quartic polynomial of week-of-the-year and a linear time (week) trend; specification 3 includes a quartic time (week) polynomial; and specification 4 includes week-of-the-year fixed effects with a linear time (week) trend.  **All models are estimated using data from 1^st^ January 2017 to 4^th^ March 2019, and model performance is assessed in the period 5^th^ March 2019 to 12^th^ August 2019. N/A: model failed to converge. | | | | | | | |

Excess deaths across all care homes for both the main analysis and selected other specifications are provided in Table A2.4. To examine robustness, we conduct four one-way sensitivity analyses where we very one element of the estimation procedure, keeping all others constant:

1. Including bed occupancy as an exposure term.
2. Allowing time trends and seasonality to be local authority-specific.
3. Model seasonality and time trends with week-of-the-year fixed effects and a linear time (week) trend.
4. Estimation using ordinary least squares.

Excess deaths from these alternative specifications are extremely close to those estimated in the main analysis.

***Table A2-4:*** *Robustness of excess death estimations to alternative econometric models*

|  | Excess deaths [95% CIs] |
| --- | --- |
| Baseline specification | 29,429 [25,047;33,810] |
| Baseline specification with bed occupancy as an exposure | 29,698 [25,400;33,996] |
| Baseline specification with local authority-specific seasonality | 29,542 [25,175;33,908] |
| Baseline specification with a linear time (week) trend and seasonality measured using week-of-the-year fixed effects | 29,717 [25,406;34,072] |
| Baseline specification estimated using ordinary least squares | 29,511 [25,150;33,873] |
| The baseline specification measures seasonality using a quartic (fourth degree) polynomial of week-of-the-year with no time trend (assuming stationarity), and is estimated using a Poisson regression with local authority fixed effects and no exposure term. All models are estimated using data from 1^st^ January 2017 to 4^th^ March 2019, and excess deaths calculated for the period 4^th^ March 2020 to 7^th^ August 2020. | |

**Appendix 3: Predicted versus observed deaths, and estimated excess deaths over time (all care homes and by care home setting type), full observational period**

***Figure A3-1:*** *Predicted versus observed deaths, and estimated excess deaths - All care homes in England. Red vertical bar marks the start of COVID-19 period in this study.*

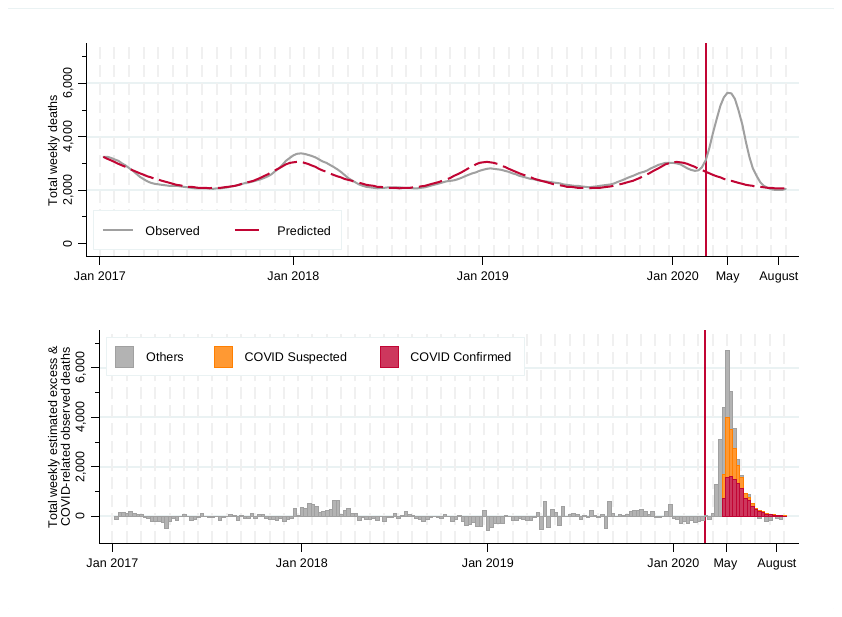

The analysis by setting type in Figure A3.2 provided further insights. Over the pre-COVID-19 period, observed weekly deaths of residents in nursing homes were of about 1.87 times (95% CI: 1.85 to 1.89) higher than in residential homes, following an almost parallel trend. In the COVID-19-period, excess deaths remained higher in nursing homes than in residential homes, by about 1.85 times but with higher variation around the weekly mean (95% CI: 1.78 to 1.92).

For both nursing and residential homes, observed deaths exceeded predicted deaths in the 2017/18 winter period. This is consistent with the moderate to high levels of influenza activity in this period, leading to a greater number of fatalities of care home residents.^[4]^

***Figure A3-2:*** *Predicted versus observed deaths, and estimated excess deaths over time by care home setting type*

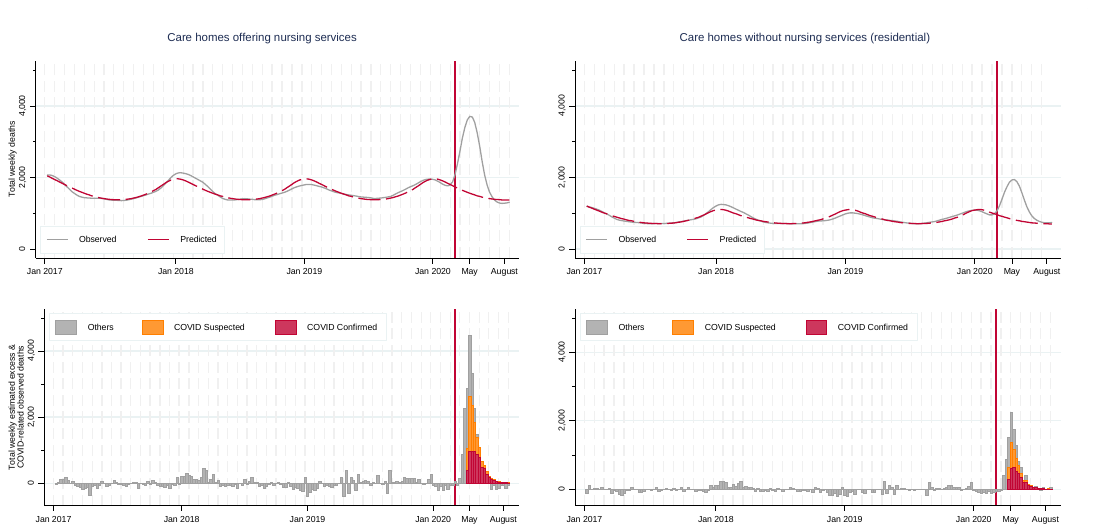

*Notes:* smoothed weekly reported death counts (observed) and predicted deaths before and after week 166 obtained from local linear regressions. **Appendix 4: Predicted versus observed deaths, and estimated excess deaths by whether care homes have experienced COVID-19 attributable deaths in the first 23 weeks of the pandemic**

**Figure A4.1** *Predicted versus observed deaths, and estimated excess deaths over time – Nursing homes*

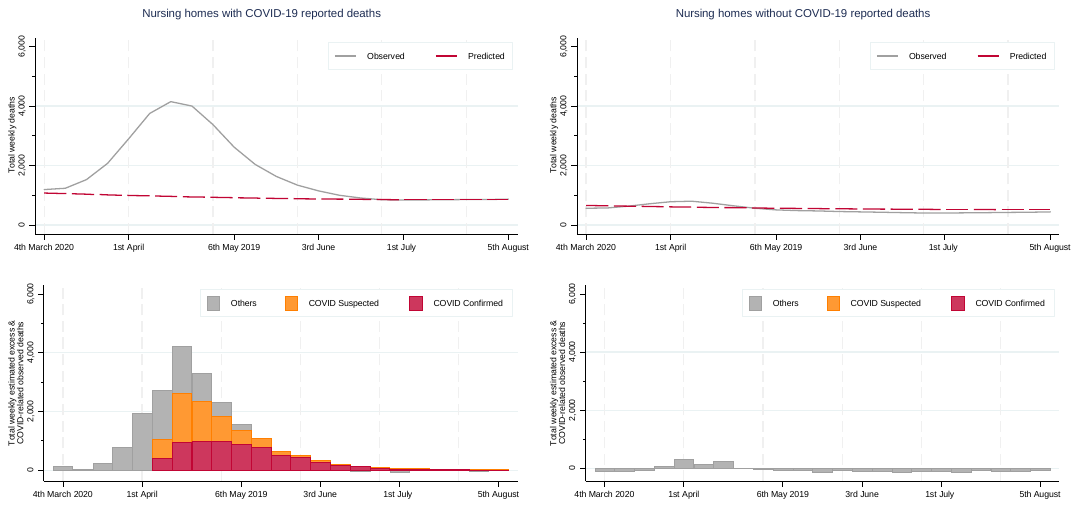

*Notes:* smoothed weekly reported death counts (observed) and predicted deaths before and after week 166 obtained from local linear regressions.

**Figure A4.2** *Predicted versus observed deaths, and estimated excess deaths – Residential homes*

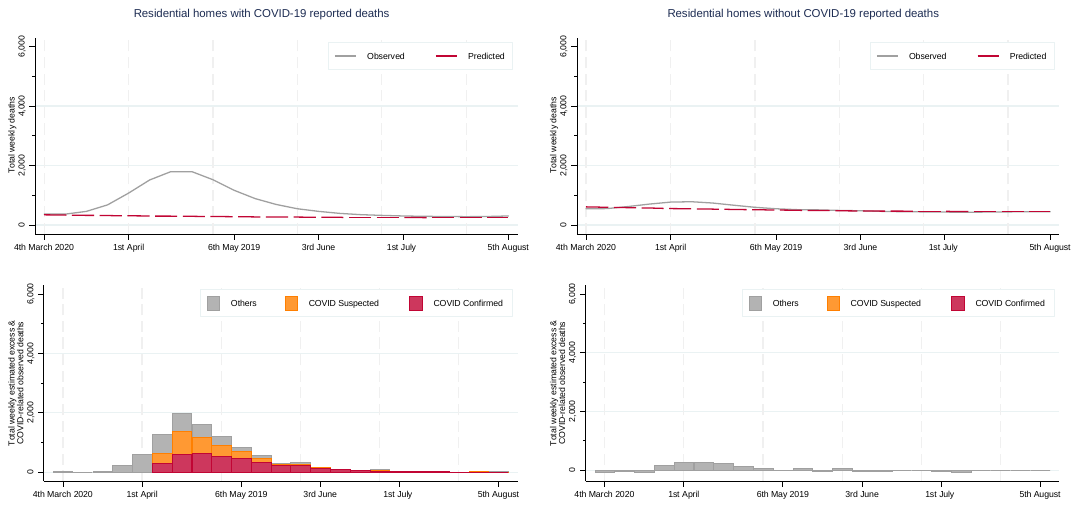

*Notes:* smoothed weekly reported death counts (observed) and predicted deaths before and after week 166 obtained from local linear regressions.

**Appendix 5: Multivariable adjusted odds ratios (sensitivity analysis)**

***Table A5-1:*** Multivariable adjusted odds ratios (95%CI) of experiencing COVID-19 confirmed/suspected deaths in the English care homes that provide services to older people/with dementia

|  | Overall | | | Nursing homes | | | Residential homes | | |
| --- | --- | --- | --- | --- | --- | --- | --- | --- | --- |
|  | OR | 95% CI | p-value | OR | 95% CI | p-value | OR | 95% CI | p-value |
| Providing nursing services | 1.75 | (1.58-1.93) | <0.001 |  |  |  |  |  |  |
| Providing residential services only | 1 | (reference) | |  |  |  |  |  |  |
| Not-for-profit care home | 0.92 | (0.79-1.08) | 0.307 | 0.87 | (0.67-1.13) | 0.292 | 0.97 | (0.8-1.17) | 0.737 |
| For-profit care homes | 1 | (reference) | | 1 | (reference) | | 1 | (reference) | |
| Branded care homes | 1.21 | (1.09-1.33) | <0.001 | 1.24 | (1.07-1.43) | 0.003 | 1.16 | (1-1.33) | 0.044 |
| Non-branded care homes | 1 | (reference) | | 1 | (reference) | | 1 | (reference) | |
| Small homes [0-23 beds] | 1 | (reference) | | 1 | (reference) | | 1 | (reference) | |
| Medium homes [24-40 beds] | 5.08 | (4.4-5.86) | <0.001 | 4.36 | (3.11-6.1) | <0.001 | 5.04 | (4.29-5.94) | <0.001 |
| Large homes [41+ beds] | 13.08 | (11.25-15.2) | <0.001 | 10.82 | (7.8-15.01) | <0.001 | 13.41 | (11.18-16.07) | <0.001 |

*Notes:* 11,134 Care homes in England providing services to older people/with dementia (4,135 nursing and 6,999 residential) reported to be active in March 2020 to CQC.

***Table A5-2:*** Multivariable adjusted odds ratios (95%CI) of an augmented model adjusted for whether care homes located in urban area (reference located in rural area) and whether in least and most deprived 20% of areas in England (2019 Index of Multiple Deprivation, IMD).

|  | Overall | | | Nursing homes | | | Residential homes | | |
| --- | --- | --- | --- | --- | --- | --- | --- | --- | --- |
|  | OR | 95% CI | p-value | OR | 95% CI | p-value | OR | 95% CI | p-value |
| Providing nursing services | 1.81 | (1.64-2) | <0.001 |  |  |  |  |  |  |
| Providing residential services only | 1 | (reference) | |  |  |  |  |  |  |
| Providing services to older people/with dementia | 5.43 | (4.34-6.79) | 0 | 2.96 | (1.97-4.46) | <0.001 | 6.56 | (4.99-8.62) | <0.001 |
| Providing non-dementia services to children and/or adults only | 1 | (reference) | | 1 | (reference) | | 1 | (reference) | |
| Not-for-profit care home | 0.96 | (0.83-1.11) | <0.001 | 0.92 | (0.72-1.17) | 0.487 | 0.99 | (0.82-1.19) | 0.905 |
| For-profit care homes | 1 | (reference) | | 1 | (reference) | | 1 | (reference) | |
| Branded care homes | 1.21 | (1.1-1.33) | <0.001 | 1.25 | (1.09-1.44) | 0.002 | 1.16 | (1.01-1.33) | 0.03 |
| Non-branded care homes | 1 | (reference) | | 1 | (reference) | | 1 | (reference) | |
| Small homes [0-23 beds] | 1 | (reference) | | 1 | (reference) | | 1 | (reference) | |
| Medium homes [24-40 beds] | 5.21 | (4.53-5.99) | <0.001 | 4.41 | (3.23-6.03) | <0.001 | 5.12 | (4.37-6.01) | <0.001 |
| Large homes [41+ beds] | 13.29 | (11.47-15.39) | <0.001 | 10.93 | (8.06-14.81) | <0.001 | 13.46 | (11.26-16.09) | <0.001 |
| Located in urban area | 1.14 | (1.01-1.29) | 0.036 | 1.21 | (1.01-1.44) | 0.038 | 1.07 | (0.91-1.27) | 0.399 |
| Located in rural area | 1 | (reference) | | 1 | (reference) | | 1 | (reference) | |
| Located in deprived area | 1.03 | (0.91-1.15) | 0.652 | 1.01 | (0.85-1.2) | 0.885 | 1.04 | (0.88-1.22) | 0.655 |
| Located in non-deprived area | 1 | (reference) | | 1 | (reference) | | 1 | (reference) | |

*Notes:* 15,524 Care homes in England (4,428 nursing and 11,096 residential) reported to be active in March 2020 to CQC.

1. Hall I, Pellis L, House T, et al. Rapid increase of Care Homes reporting outbreaks a sign of eventual substantial disease burden. *medRxiv* 2020:2020.05.07.20089243. doi: 10.1101/2020.05.07.20089243
2. Cox NJ. Stata tip 111: More on working with weeks. *The Stata Journal* 2012;12(3):565-69.
3. Competition Markets Authority. Care homes market study,: Competition and Markets Authority,

   London https://assets.publishing.service.gov.uk/media/5a1fdf30e5274a750b82533a/carehomes-

   market-study-final-report.pdf, 2017.
   Espuny Pujol F, Hancock R, Hviid M, et al. Market concentration, supply, quality and prices paid

   by Local Authorities in the English care home market: Centre for Competition Policy,

   University of East Anglia, Norwich, UK., 2019 (under revision in Health Economics).
4. Public Health England (2018) Surveillance of influenza and other respiratory viruses in the UK: Winter 2017 to 2018. https://assets.publishing.service.gov.uk/government/uploads/system/uploads/attachment_data/file/740606/Surveillance_of_influenza_and_other_respiratory_viruses_in_the_UK_2017_to_2018.pdf>
